# SPELL: A Scalable NLP Method Using Regular Expressions and Large Language Models for Clinical Information Extraction

**DOI:** 10.1101/2025.07.25.25332130

**Authors:** Ricardo Kleinlein, David W. Bates, Carolyn Guan, Kathryn J. Gray, Vesela P. Kovacheva

## Abstract

**Objective:** Electronic health records (EHRs) contain valuable information for clinical research and decision-making. However, leveraging these data remains challenging due to data heterogeneity, inconsistent documentation, missing information, and evolving terminology, especially within unstructured clinical notes. We developed SPELL (**S**nippet-**P**rimed r**E**gex **LL**M Pipeline), a scalable natural language processing (NLP) workflow to systematically extract structured clinical insights from large volumes of clinical narratives.

**Materials and Methods:** Our platform employs a hybrid approach combining regular expressions (regex) to rapidly identify relevant textual snippets with locally hosted large language models (LLMs) for accurate clinical interpretation. All data processing occurs securely within institutional computational environments. The modular Python-based workflow facilitates adaptation across institutions and is optimized for computational efficiency, supporting high-throughput processing even in resource-limited settings. We quantified computational scalability (elapsed time, out-of-memory events, GPU temperature, and energy consumed) and audited retrieval recall using clinician-annotated regex-negative notes enriched with relevant structured metadata.

**Results:** The pipeline efficiently processed 31 million clinical reports spanning 1976–2024 from eight affiliated hospitals. By analyzing targeted snippets rather than entire documents, our approach reduced processing time by 68% compared to traditional full-document LLM inference, and by >95% compared to manual physician annotation. Accuracy was rigorously validated across three obstetric tasks: extraction of numerical values (blood loss volumes), dates (estimated due dates), and diagnoses (hemolysis, elevated liver enzymes, and low platelets [HELLP] syndrome). Task-level performance included 94-98% exact-match accuracy for the three concepts on curated snippets. Generalizability was investigated using the publicly available MT Samples corpus (5,013 notes, 40 specialties), yielding 84% accuracy for ventricular tachycardia detection with markedly fewer false positives.

**Discussion and Conclusions:** A hybrid regex→snippet→LLM approach delivers accurate, privacy-preserving, and computationally efficient extraction for unstructured EHR data. By focusing inference on snippets and deploying local, open-weights models, SPELL aligns with institutional data governance requirements while enabling scalable clinical informatics studies across diverse extraction tasks.

**Summary Statement:** We developed SPELL, a scalable NLP pipeline combining regex and locally hosted LLMs for efficient information extraction from clinical narratives.

## INTRODUCTION

Electronic health records (EHR) contain extensive clinical information, stored both as structured data (e.g., laboratory results, medications) and unstructured narrative text (e.g., physician notes, imaging reports). While structured data can be readily analyzed computationally, unstructured narratives often contain critical clinical details not captured elsewhere [1]. Extracting useful insights from clinical narratives can significantly enhance clinical decision-making [2], research [3,4], and predictive analytics [1,5]. However, using unstructured EHR data remains challenging due to heterogeneity, inconsistent documentation, and the complexity inherent in clinical language [6,7]. Currently, manual annotation is the de facto standard method for extracting clinical information from text, but it is labor-intensive [8,9], resource-consuming [9,10], and subject to human annotator variability [11], limiting large-scale or longitudinal studies [10]. Therefore, there is a clear need for automated natural language processing (NLP) methods capable of accurately and efficiently extracting structured clinical data from unstructured text.

Early clinical NLP systems primarily relied on rule-based and statistical methods, which require significant manual feature engineering, have limited generalizability, and necessitate frequent retraining as clinical documentation evolves [8,12]. Recently, transformer-based large language models (LLMs), capable of zero- or few-shot inference, have emerged as powerful alternatives. These models can substantially reduce annotation burdens, better capture contextual nuances, and demonstrate broad adaptability across diverse clinical extraction tasks [13–15]. Nonetheless, many existing LLM-based clinical NLP frameworks often rely on cloud-based systems, which may pose challenges regarding institutional data governance policies, the handling of protected health information, and high operational costs [13–17].

Hybrid approaches combining regular expression (regex)-based preprocessing with LLM inference have been explored [14,18]. However, existing hybrid pipelines frequently lack systematic validation of regex retrieval performance, detailed measures of computational scalability, and rigorous comparisons with manual annotation. Additionally, prior approaches often perform inference at the full-document level, incurring high computational costs, or rely exclusively on cloud-hosted models [14,18]. Thus, there remains a strong rationale for a systematic, computationally efficient hybrid NLP pipeline explicitly designed for institutional clinical deployments.

To address these gaps, we developed SPELL (**S**nippet-**P**rimed r**E**gex **LL**M Pipeline), a scalable NLP workflow combining regex-based snippet extraction with locally deployed LLM inference.

Scalability, as defined in this manuscript, explicitly refers to the capability of efficiently processing large-scale EHR datasets (on the order of millions of clinical documents), with predictable computational resource utilization and inference times. SPELL significantly reduces computational load by restricting inference to targeted textual snippets rather than entire documents. We quantify scalability through computational metrics, including graphics processing unit (GPU) temperature, total processing elapsed time, GPU energy consumption per process, and out-of-memory events, which are rarely reported in prior hybrid (regex-plus-LLM) NLP evaluations.

We demonstrate the utility and generalizability of SPELL through rigorous evaluation across three representative obstetric clinical tasks selected due to their frequent reliance on narrative documentation and limited representation in structured data: quantifying blood loss (BL) volumes, extracting estimated due dates (EDD), and identifying diagnoses of hemolysis, elevated liver enzymes, and low platelets (HELLP) syndrome. Methodologically, we emphasize clear evaluation standards, nonparametric statistical uncertainty quantification, systematic regex retrieval auditing, and detailed characterization of computational scalability. To further assess generalizability beyond our institutional data, we evaluate SPELL on the publicly available MT Samples dataset[19], specifically evaluating its ability to detect ventricular tachycardia diagnoses across diverse clinical notes.

In summary, SPELL represents a deployment-ready, computationally optimized NLP pipeline designed explicitly for institutional-scale clinical narrative processing, addressing key limitations of existing hybrid NLP methods and significantly advancing the feasibility of automated clinical information extraction at scale.

## 2. BACKGROUND AND SIGNIFICANCE

Early clinical NLP systems relied on manually crafted, rule-based methods, which were effective primarily for numeric data and structured entities, but were labor-intensive and lacked flexibility for diverse clinical documentation [2,12,20,21]. The emergence of transformer-based language models (e.g., BioBERT, ClinicalBERT) significantly advanced clinical text analysis through enhanced context-aware interpretation [22–25]. However, these models require extensive annotated corpora and task-specific fine-tuning, limiting practical scalability in clinical workflows [22].

Generative LLMs enable zero-/few-shot information extraction and can reduce annotation effort, though performance is task- and domain-dependent [13,14,26,27]. Many reported deployments use cloud services, while on-premises options are increasingly available; in practice, hosting choice is a governance decision rather than a methodological one. Ontology-driven tools (e.g., cTAKES, MetaMap) integrate standard terminologies (UMLS, SNOMED CT, ICD) and provide robust concept mapping, yet they may miss rare or context-dependent mentions and typically require add-on modules for negation and temporality, increasing operational complexity [4,28–30].

Recent studies have combined regex-based pre-processing with generative LLMs for clinical data extraction. For example, one recent study integrated regular expressions with GPT-4 to extract surgical data but provided limited systematic validation of regex extraction performance, particularly regarding precision, recall, or error categorization [18]. Another evaluation of open-source LLMs for extracting structured social determinants of health (SDoH) data demonstrated improved performance compared to traditional regex methods [14]. However, this method required task-specific prompt refinement and pipeline customization and was evaluated primarily on a single health system and set of SDoH concepts, without explicitly demonstrating rapid adaptability or generalization across diverse clinical extraction scenarios [14]. Similarly, a clinical entity-augmented retrieval method demonstrated computational efficiency gains by leveraging clinical entities for information retrieval, yet did not quantify manual annotation burden or provide explicit evaluation across a range of clinical specialties [31].

SPELL explicitly addresses these methodological gaps by systematically validating regex snippet-based retrieval performance, introducing a snippet-primed inference method to substantially reduce computational load, achieving substantial clinician annotation and computational efficiency gains, and demonstrating zero-shot adaptability and initial generalizability across multiple clinical domains, supported by detailed computational benchmarks (GPU throughput, dynamic scheduling, energy consumption).

## 3. MATERIALS AND METHODS

### 3.1 Ethics and Data Governance

All research activities adhered to institutional policies governing data privacy, ethical standards, and data governance, and received approval by the Mass General Brigham Institutional Review Board (IRB#2020P002859). All clinical data were processed on-premises within an institutional computational environment following institutional data governance requirements. Data were handled exclusively in pseudonymized form, with personally identifiable information such as patient names, medical record numbers, Social Security Numbers, contact details, and precise dates systematically obfuscated or entirely removed.

### 3.2. Data Sources, Indexing, and Cohort Selection

#### 3.2.1. Clinical Data Sources and Indexing

Clinical narratives were sourced from a centralized institutional data warehouse aggregating EHR across multiple affiliated hospitals [32]. Clinical notes were received as plain text files containing multiple reports, each delimited by standardized headers and end-of-report tokens. To optimize storage and accelerate retrieval, we developed a unified parsing method for these files and created byte-offset indexes keyed by patient pseudo-ID, encounter ID, note type, and timestamp, without duplicating note content (Figure 1). This indexing strategy improved retrieval efficiency by storing byte offsets and document lengths rather than duplicating notes, which significantly reduced storage and accelerated retrieval.

**Figure 1.**
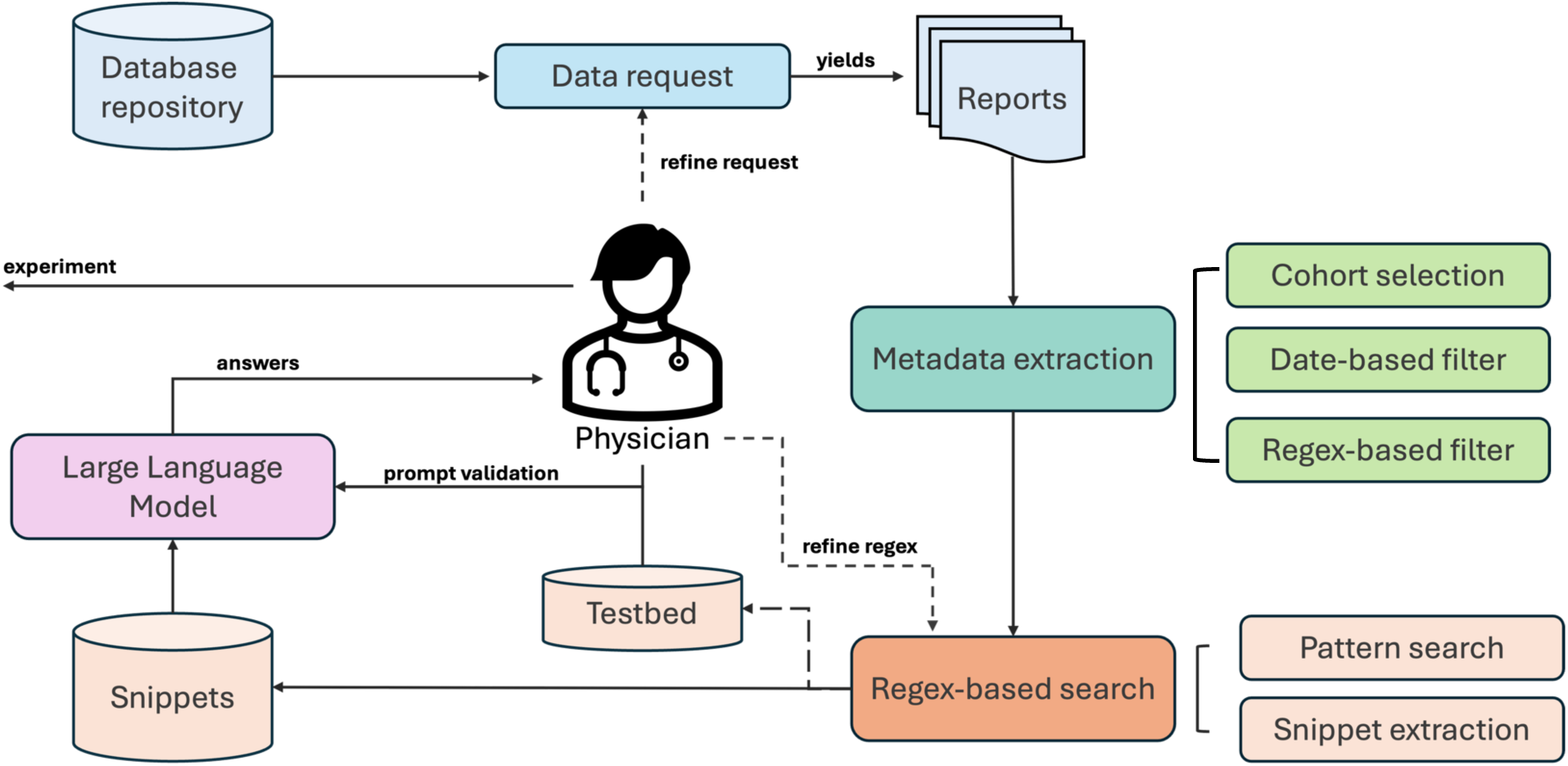
Overview of the SPELL platform. The system comprises four key components: (1) Data request—clinicians and researchers retrieve clinical data from institutional database repositories; (2) Metadata extraction—a preprocessing step that indexes reports and extracts key metadata to streamline cohort selection; (3) Regex-based search—a filtering module that eliminates irrelevant reports and extracts snippets from relevant matches; and (4) large language model (LLM)-based retrieval, a locally hosted LLM that analyzes extracted snippets to enhance information retrieval efficiency. This iterative workflow enables precise alignment between selected patient cohorts and research objectives.

#### 3.2.2. Cohort Definition and Filtering

Study cohorts were precisely defined by demographic attributes, clinical conditions, note types, and specific temporal windows (e.g., gestational periods). Data selection was conducted using structured metadata queries combined with regular expression (regex)-based textual searches. Domain-specific regex patterns were collaboratively developed by clinicians and data scientists and iteratively refined via random sampling quality assessments (Table 1).

**Table 1:**
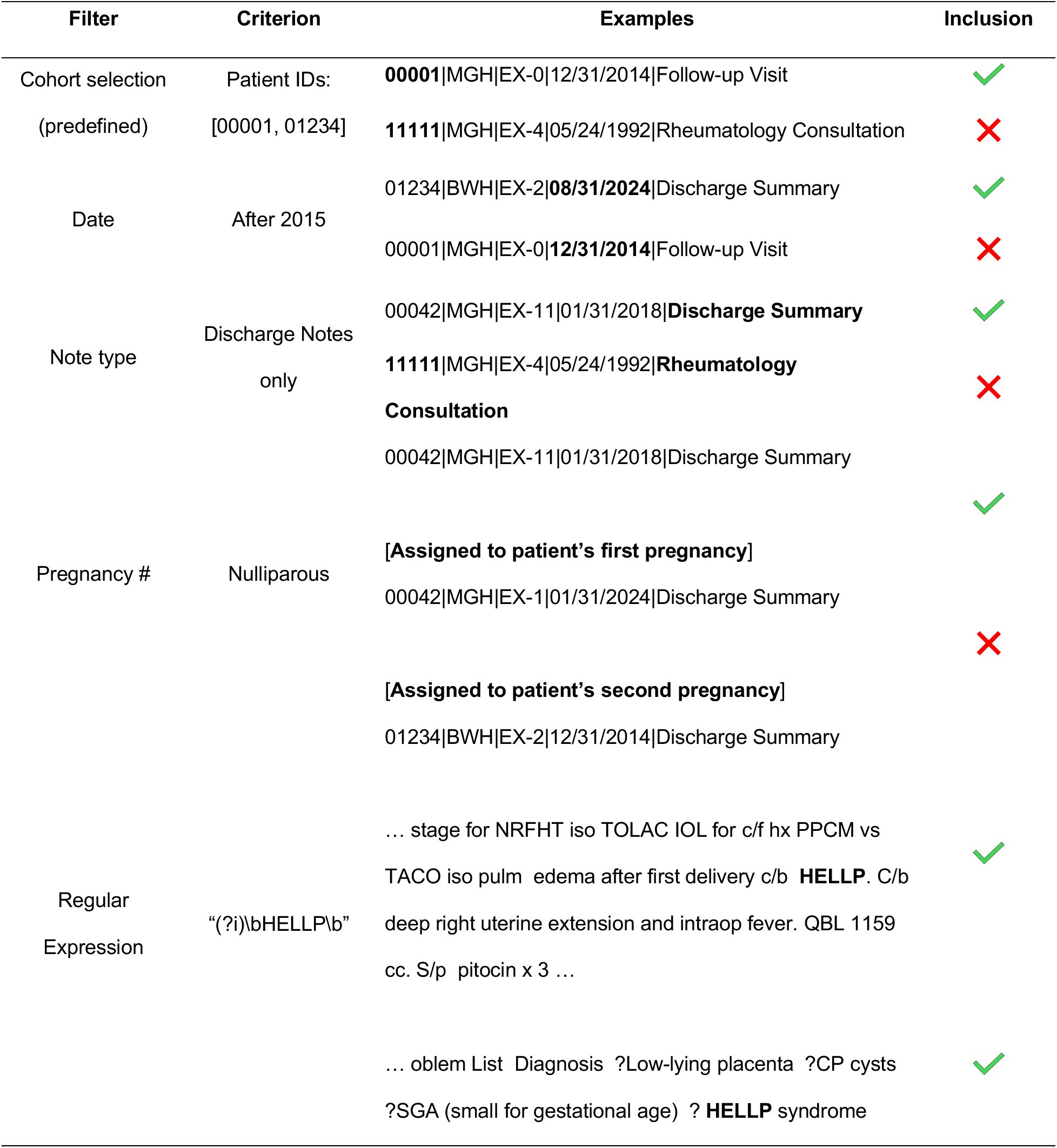
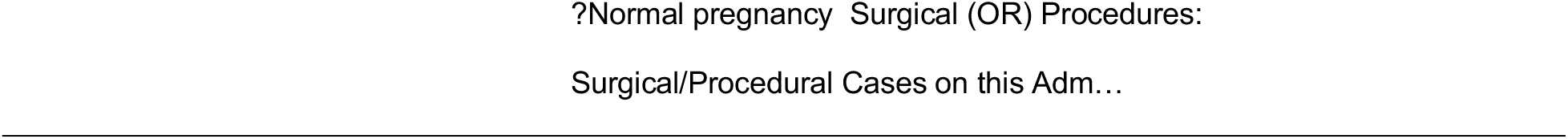
Examples of filters implemented in the dataset generation process.

#### 3.2.3. Formal Problem Setup

The information extraction task was formally defined using a two-stage retrieval–inference approach. Given the evaluation set 𝒟 containing *N* clinical notes, each document *d* ∈ 𝒟 underwent retrieval followed by inference.

In the retrieval phase, a regex-based retrieval function *R*_Θ_(*d*), parameterized by predefined patterns Θ = {θ_1_, θ_2_, … , θ_3_} and a character window *w*, returns *k*_*d*_ ≥ 0 snippets 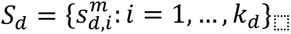 around the matches found for every θ_+_ (with *m* ≤ *M*) of the predefined patterns, given that ∃*θ*_*m*_ ∈ Θ, ∃ *s*_*d*,*i*_ ⊆ *content*(*d*): *s*_*d*,*i*_ ⊨ *θ*_*m*_. In the inference phase, the snippets extracted by *R*_Θ_(*d*) are inputted to a task-prompted, pretrained LLM Φ, generating document-level inferences:

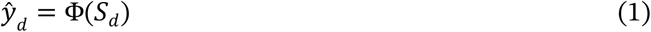

Clinician-annotated labels *y*_*d*_ served as the gold standard for evaluation, varying by task. For BL extraction, annotations consisted of numeric values in milliliters (mL); for EDD, annotations were ordinal days relative to a canonical reference date; and for HELLP syndrome or ventricular tachycardia detection, annotations were binary, indicating presence or absence within the note. A canonicalization function g(⋅) normalized LLM outputs into comparable numeric or categorical forms for direct comparison with clinician annotations. If the model returned “None,” we set *ŷ*_%_ = Ø. Since retrieval acted as a gate for inference, the overall pipeline sensitivity can be expressed as:

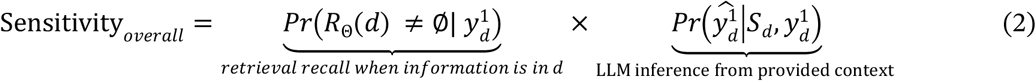

Where 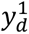 denotes that the information sought to extract in document *d* is present. Retrieval recall was explicitly audited through random sampling to quantify false negatives from regex retrieval.

### 3.3. Pipeline Overview and Information Extraction Workflow

The extraction pipeline involved two sequential stages (Figures 1–2). First, clinical notes were searched using task-specific regex patterns, such as “qbl|ebl|blood\s*loss” for BL, “edd|estimated date of delivery” for EDD, and “\bH\.?E\.?L\.?L\.?P\b” for HELLP syndrome detection. Snippets surrounding regex matches were extracted with a default ±100-character window. It is important to highlight that even though the window length around every match is constant, the number of matches can vary for every document.

**Figure 2.**
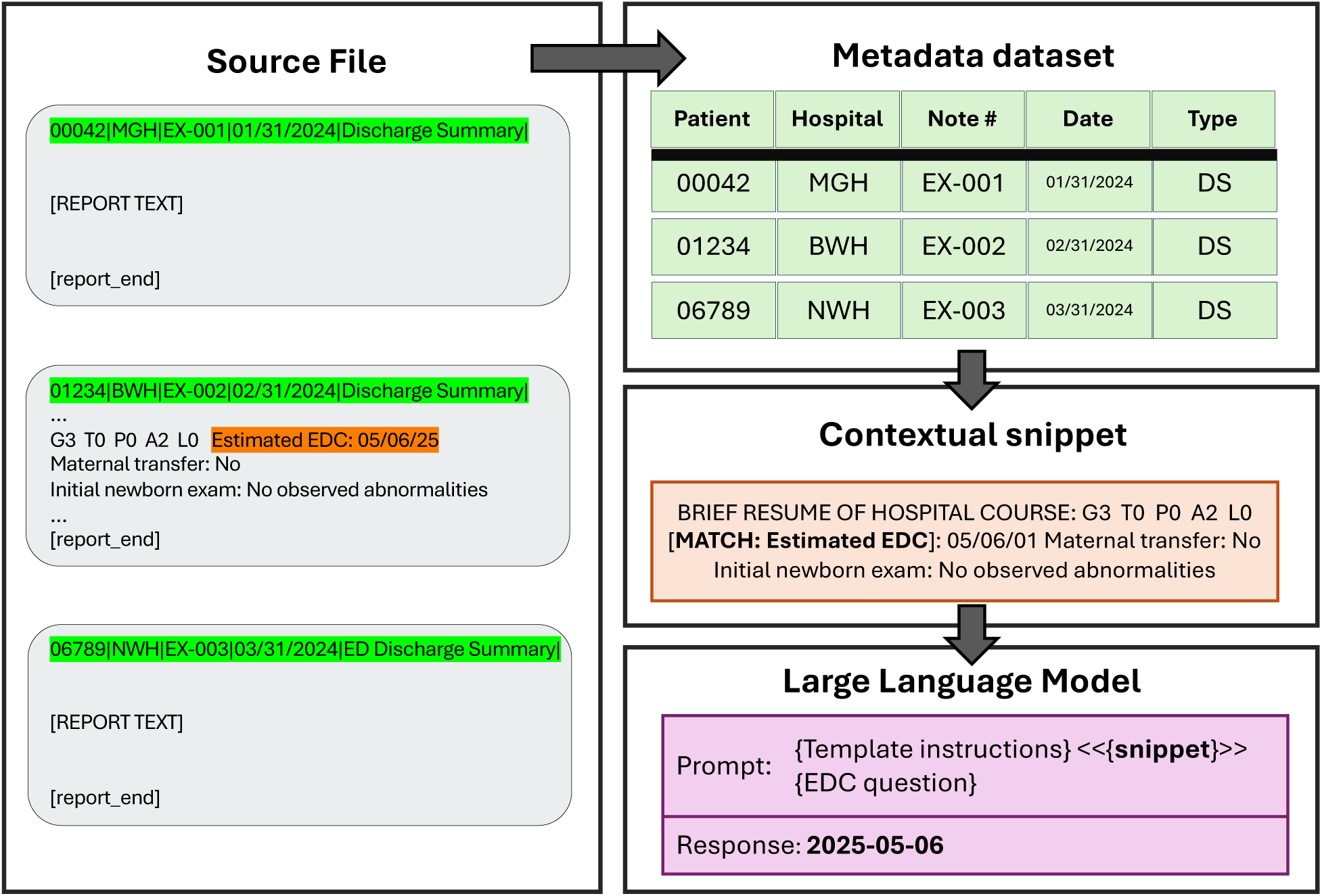
Overview of the system workflow in SPELL, illustrating the functionality of each module through a representative example present in the estimated due date (EDD) extraction task. Each block demonstrates a specific module in action, with arrows indicating the sequential processing steps from dataset creation to information extraction.

In the second stage, snippets were analyzed by a locally hosted pretrained LLM (Llama 3.1-8B-Instruct), guided by task-specific prompts. These predictions were post-processed into canonical forms (e.g., numeric units, standardized dates) for evaluation. Given inefficiencies from padding variable-length texts, we employed a single-sample dynamic scheduling strategy processing each snippet individually, substantially reducing computational overhead and improving throughput. Specifically, let *T*(•) denote the time required by the LLM to process an input. We define padding overhead ratio (*ρ*_pad_) as:

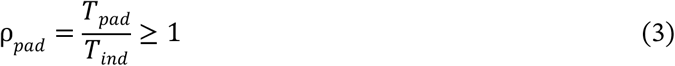

Where

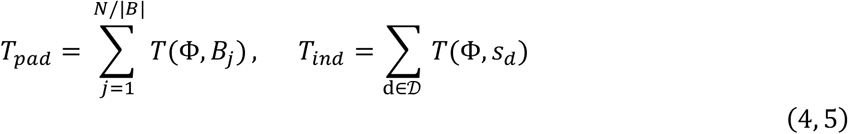

Where *B*_*j*_ represents a batch of snippets 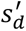 where each of these document snippets is zero-padded to match the length of the longest snippet in the batch. Our scheduler minimized this padding-induced overhead. Computational efficiency was further quantified via energy consumption metrics (Appendix S1):

#### 3.3.1. Regex-guided Snippet Retrieval

Relevant notes were identified using regex patterns collaboratively defined by clinical experts (V. K. and K.G.). Patterns targeted specific clinical terms (e.g., BL: “qbl|ebl|blood\s*loss”, EDD: “edd|estimated date of delivery”, HELLP: “\bH\.?E\.?L\.?L\.?P\b”). Matched text was extracted with ±100-character context windows, adjusted as needed for each task. Regex patterns were tested across multiple note types (discharge summaries, History&Physical, operative reports) from eight affiliated hospitals to ensure consistent retrieval behavior despite documentation heterogeneity.

#### 3.3.2. LLM-based Information Extraction

Extracted snippets were analyzed using a locally hosted generative LLM (Llama 3.1-8B-Instruct), an autoregressive transformer instruction-tuned via supervised fine-tuning and reinforcement learning with human feedback. We selected the 8-billion-parameter conversational Llama model primarily because its moderate size allowed for efficient, high-throughput inference on standard institutional GPU hardware, while offering a favorable accuracy–efficiency trade-off. Recent evidence suggests that instruction-tuned generative LLMs, including Llama-family models, can achieve performance comparable to state-of-the-art closed-source LLMs (e.g., GPT-4o) and outperform traditional transformer-based NLP models (e.g., BERT) in clinical information extraction tasks, particularly under zero-shot conditions or when only modest fine-tuning data are available [17]. However, our pipeline is designed to be model agnostic, hence offering flexibility to employ other state-of-the-art LLMs.

The pipeline is best suited for use with modern LLMs, which, as a rule, have context windows exceeding several thousand tokens, a length our targeted snippets rarely meet. However, to ensure computational efficiency while maintaining contextual coherence, snippets exceeding context window limits were segmented using a 50% overlapping sliding-window approach. The LLM-generated outputs were obtained using explicitly configurable inference parameters shared across documents. These include generation temperature (*T*, maximum response token length (*L_max_*), and task-specific prompt templates (*P*). The clinical information extraction was then formally defined as:

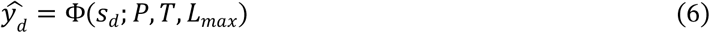

We carefully developed task-specific prompts (Appendix S8), instructing the model explicitly to return only the requested clinical value or label. For example, the estimated due date (EDD) extraction required ISO-standard dates, while the BL extraction required numeric values in milliliters. Default decoding parameters included deterministic decoding (temperature = 0.0, top_p = 1.0), a maximum of 3 generated tokens for the BL and HELLP extraction and 7 for the EDD task, and a fixed random seed (42) for reproducibility across runs. Final outputs underwent rigorous post-processing for numeric parsing, unit normalization, and strict date-format validation according to the annotation guidelines (Section 3.4.1).

#### 3.3.3. Computational Optimizations: Chunking and Scheduling

To address computational inefficiencies arising from padding variable-length inputs, we implemented dynamic single-sample scheduling strategy. Snippets exceeding the model’s maximum token limit were segmented into smaller chunks, using a sliding-window approach with 50% overlap to maintain contextual continuity and avoid truncation.

The platform allowed inference parameters—including generation temperature, maximum token length, and prompt templates—to be adjusted flexibly between tasks, enabling optimization for computational efficiency and extraction accuracy. Inference was intentionally restricted to targeted, regex-identified snippets rather than complete clinical documents, aiming to reduce computational load and minimize irrelevant textual input. Task-specific prompts were explicitly crafted to facilitate accurate clinical information extraction without requiring additional model fine-tuning or extensive prompt engineering (see Appendix S8). All inference parameters, prompts, and scripts were systematically documented to ensure reproducibility and enable iterative refinement.

### 3.4. Annotation Protocol and Evaluation Strategy

#### 3.4.1. Annotation Protocol

We developed concise annotation guidelines, accompanied by clinical examples, to ensure clarity and consistency. For BL, annotators extracted total quantitative or estimated blood loss values in milliliters, prioritizing explicitly stated totals when multiple values appeared, and normalized units (e.g., “1 L” → 1000 mL). For EDD, annotators extracted a single date in ISO standard format (YYYY-MM-DD), choosing the latest clinician-asserted date recorded prior to delivery if multiple dates appeared, otherwise returning “None.” For binary classifications (HELLP/ventricular tachycardia), notes were annotated positive only if the diagnosis or suspicion was explicitly confirmed for the current encounter or pregnancy; historical, ruled-out, or uncertain mentions were annotated negative.

Clinical annotations were independently performed by two clinicians (V.K. and C.G.), with discrepancies resolved through adjudication by a third clinical expert (K.G.). Inter-annotator reliability was assessed separately for numeric extraction tasks (BL, EDD) using exact agreement rates (percentage of cases in which annotators provided identical annotations), and for the binary classification task (HELLP syndrome) using Cohen’s κ statistic.

#### 3.4.2. Evaluation Metrics

The evaluation employed document-level analysis. Strict exact match was formally defined as:

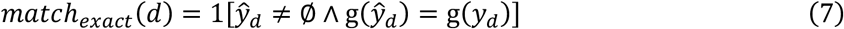

Where *g*(•) denotes the canonicalization function described above. Binary classification metrics, including Precision (P), Recall (R), F₁-score, and Accuracy, were computed using standard definitions with micro-averaging:

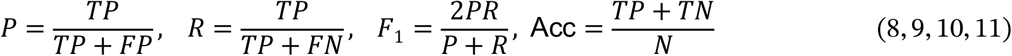

As a comparator, we implemented a word-window regex-only baseline (denoted Regex@N). For each regex match, we extract the first candidate within N whitespace-delimited words after the anchor inside a snippet. We report on exact matching accuracies for N=1, 3, and 5, corresponding to 1, 3, and 5-word windows.

To quantify uncertainty, we computed 95% confidence intervals using nonparametric bootstrap resampling over documents (B=1000, percentile method). Paired method comparisons between LLM and regex extraction with a 5-word window used McNemar’s test for binary tasks:

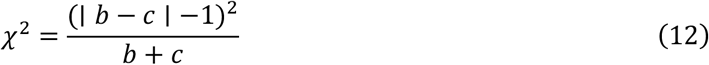

with *b* denoting the number of cases for which the LLM is correct but the Regex baseline is incorrect, and *c* represents the number of instances for which the Regex baseline is correct but the LLM is incorrect.

#### 3.4.3. Retrieval Recall Audit

Given the rarity of certain clinical events (e.g., HELLP syndrome, severe blood loss), a purely random sampling approach was not feasible due to an expected very low positivity rate. Instead, we conducted a targeted recall audit using notes with no regex matches but with supportive clinical metadata (e.g., ICD codes, laboratory values) suggestive of relevant clinical content. A clinician independently annotated these targeted notes, identifying true positives that were missed by regex-based retrieval. We explicitly computed retrieval recall on this enriched subset as:

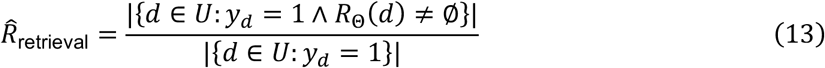

where *U* is a clinician-validated, enriched subset rather than a random sample of all notes. Accordingly, *R̂*_retrieval_ reflects retrieval performance within this enriched frame and may overestimate recall in the full corpus.

#### 3.4.4. Human-with-Snippets Baseline

To isolate the specific advantage of snippet-based retrieval, we had clinicians independently annotate identical snippet sets presented to the LLM for a representative subset of 50 notes per task. Annotation accuracy and timing metrics from snippet-based human annotation were directly compared to annotations obtained from full-document reviews and the LLM scenario.

### 3.5. Computational Efficiency and Metrics

We explicitly quantified computational performance by defining total tokens processed in both full-document (*T*_full_) and snippet-based (*T*_snip_) scenarios as:

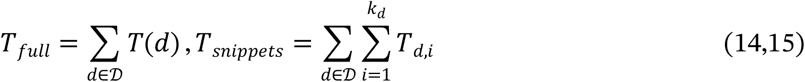

Computational speedup attributable to snippet-based inference was then reported as:

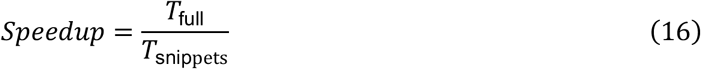

excluding constant I/O and scheduling overhead. Throughput was documented as elapsed time. ‘Elapsed time’ refers to end-to-end time measured from job start to completion, including tokenization and inference (excluding indexing), unless otherwise specified. GPU energy consumption was measured from power logs alongside GPU temperature monitoring (Appendix S1).

### 3.6. Clinical Use Cases and External Validation

Performance was rigorously validated on three obstetric clinical extraction tasks (BL, EDD, HELLP) using clinician annotations as the gold standard. To further assess generalizability, the pipeline was evaluated externally on ventricular tachycardia detection using the MT Samples dataset, consisting of 5,013 notes across 40 clinical specialties[19].

### 3.7. Hardware and Software Infrastructure

The pipeline was developed and executed on a single Linux workstation (Ubuntu Linux v22.04 LTS) equipped with three NVIDIA RTX A4000 GPUs, each with 16 GB of VRAM. Computational environments were managed using a dedicated Miniforge installation (Python v3.13.5, CUDA v12.7), with essential computational libraries including Polars (v1.17.1) for efficient tabular data manipulation and Transformers (v4.45.2) for local LLM inference.

## 4. RESULTS

### 4.1. Corpus Overview and Data Indexing

We processed 149 GB of unstructured EHR narratives encompassing 30,888,929 clinical reports from 242,413 unique patients across 8 affiliated hospitals from 1976–2024 (Appendices S2–S5). Metadata indexing completed in 85 minutes, generating 3.15 GB of byte-offset indices to streamline retrieval. The unit of analysis was the individual clinical document.

### 4.2. Regex-based Note Selection and Retrieval Coverage

#### 4.2.1. Regex Retrieval Performance

Clinical notes relevant to the three obstetric tasks were identified using collaboratively designed regex patterns (Appendix S6) with ±100-character windows around matched text. Retrieval times were 70–104 minutes, yielding initial candidate pools of 1,286,161 notes for BL (∼104 minutes), 2,066,023 notes for EDD (∼81 minutes), and 46,953 notes for HELLP syndrome (∼70 minutes).

Final evaluation subsets selected for downstream annotation and performance assessment comprised 92,380 notes (BL), 35,172 notes (EDD), and 540 notes (HELLP), as detailed in Appendix S7.

#### 4.2.2. Retrieval Coverage and Recall Audit

Document-level retrieval recall (*R̂*_retrieval_) was independently audited via a regex-independent sample (n = 50/task). In our regex recall audit, we reviewed 50 regex-negative notes per task (BL, EDD, and HELLP syndrome), explicitly selected using structured metadata indicative of potential clinical relevance. No missed positive cases (false negatives) were identified for BL, EDD, or HELLP syndrome, resulting in an estimated recall of 1.00 for all three tasks. Given the small sample size and non-random sampling frame, the true recall in the full corpus may be lower.

### 4.3. Computational Efficiency and Scalability

#### 4.3.1. Annotation Time and Speedup Comparisons

We benchmarked the LLM-based snippet approach (Llama 3.1-8B-Instruct, deterministic decoding) against manual clinician review and LLM full-document processing. Table 2 summarizes the mean per-document annotation times, standard deviations, and relative speedups comparing manual clinician review and LLM processing approaches. LLM times report means ± standard deviation (SD) from 100 replicates.

**Table 2:** Annotation times (in seconds) comparing physician with LLM extraction. Physician annotation times are reported as means ± standard deviation (SD) across the two annotating clinicians (V.K. and C.G.), and LLM times are reported as means ± SD across 100 repeated runs.

| Use case<br>(time in s) | Physician<br>(snippets) | LLM<br>(snippets) | Physician<br>(full document) | LLM<br>(full document) |
| --- | --- | --- | --- | --- |
| Blood loss | 396.5 (52.5) | 12.65 (0.45) | 1172.5 (28.5) | 73.71 (0.36) |
| EDD | 459 (92.0) | 19.54 (0.16) | 1874.5 (368.5) | 66.27 (0.17) |
| HELLP | 440 (40.0) | 10.59 (0.16) | 1942.0 (268.0) | 79.96 (0.35) |

Abbreviations: EDD, estimated due date; HELLP, hemolysis, elevated liver enzymes, low platelets, LLM large language model.

#### 4.3.2. Dynamic Scheduling

Dynamic single-sample scheduling strategy reduced the total elapsed time required to process a set of 540 clinical notes by up to 76% (*ρ*_*pad*_ = 4.31) compared to traditional fixed-length batch processing (Fig. 3), aligning with the high variability in snippet lengths (12–2,000 tokens). Total tokens processed using snippet-based inference were ∼22.8 million tokens across the three tasks (BL: 19.38M; EDD: 3.34M; HELLP: 0.084M).

**Figure 3:**
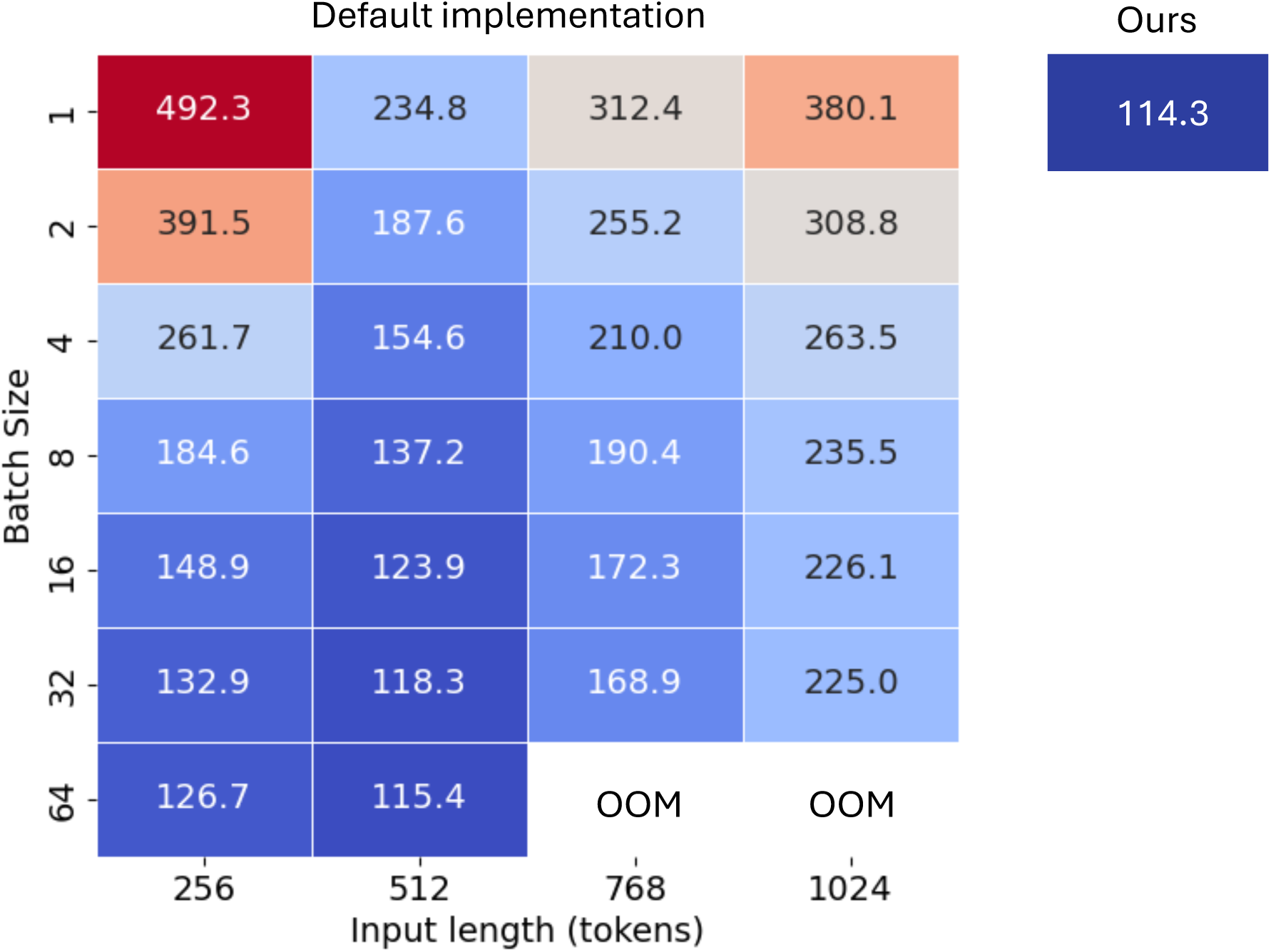
Total processing time (in seconds) for 540 clinical notes across different batch sizes (y-axis) and input lengths in tokens (x-axis) using the preferred implementation of the Hugging Face Transformer pipeline. Each cell represents the total runtime for a specific combination of these hyperparameters. Two configurations resulted in Out-Of-Memory (OOM) errors. The right panel displays our dynamic, single-sample scheduler total runtime on the same dataset.

### 4.4. Document-level Extraction Accuracy and Reliability

Performance metrics include strict matching for BL and EDD, and standard classification metrics for HELLP. Regex-only extraction (Regex@N; N ∈ {1,3,5}) served as a baseline.

Annotation reliability was high across tasks. For numeric extraction tasks, exact agreement rates between annotators (V.K. and C.G.) were 88% for full documents and 96% for snippets (BL extraction), and 92% for full documents and 86% for snippets (EDD extraction). For the binary classification task (HELLP syndrome), Cohen’s κ was 0.786 (full documents) and 0.847 (snippets), indicating strong agreement. These annotated data served as the gold standard for evaluating the accuracy of the LLM-assisted extraction (Table 3).

**Table 3.**
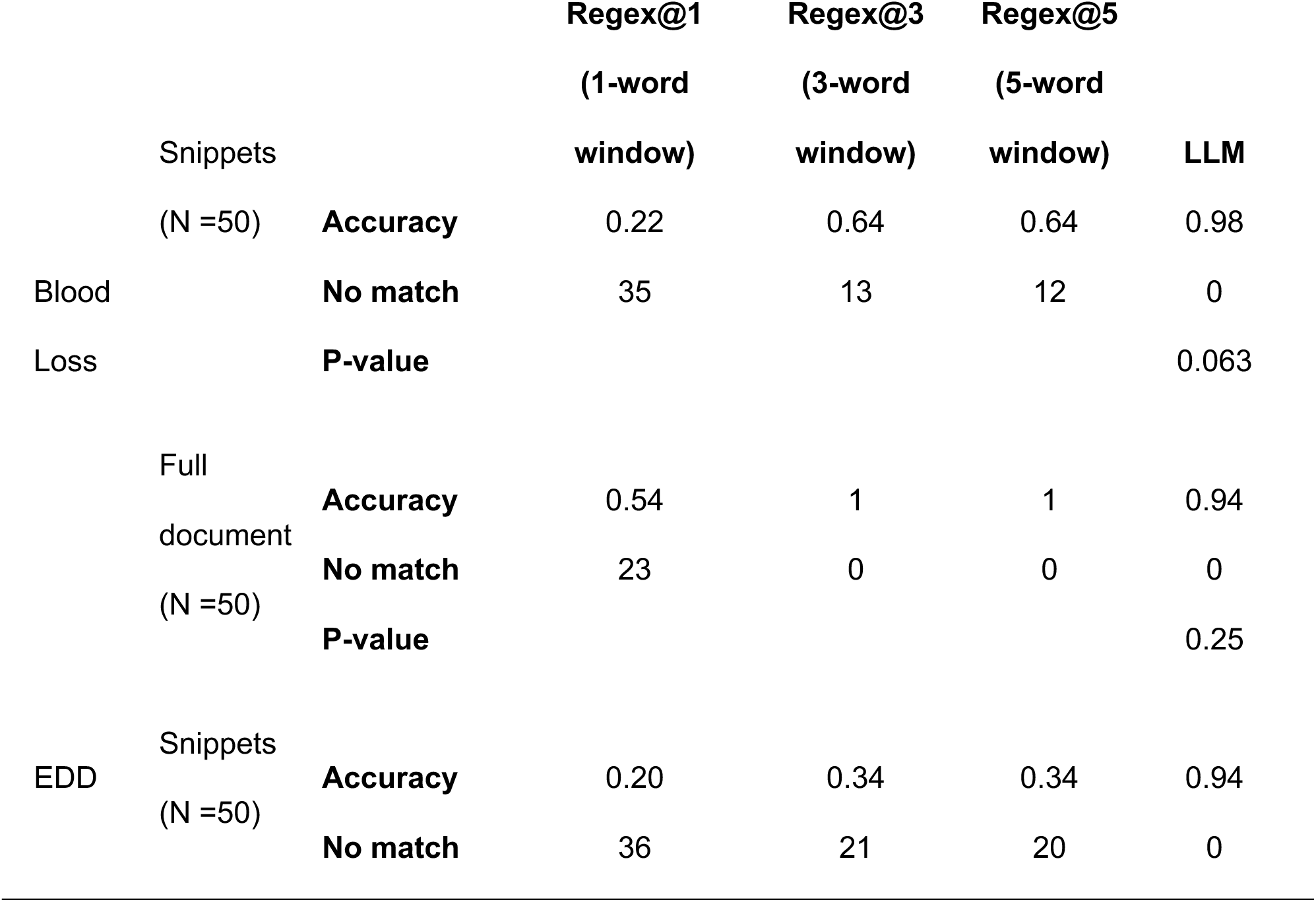
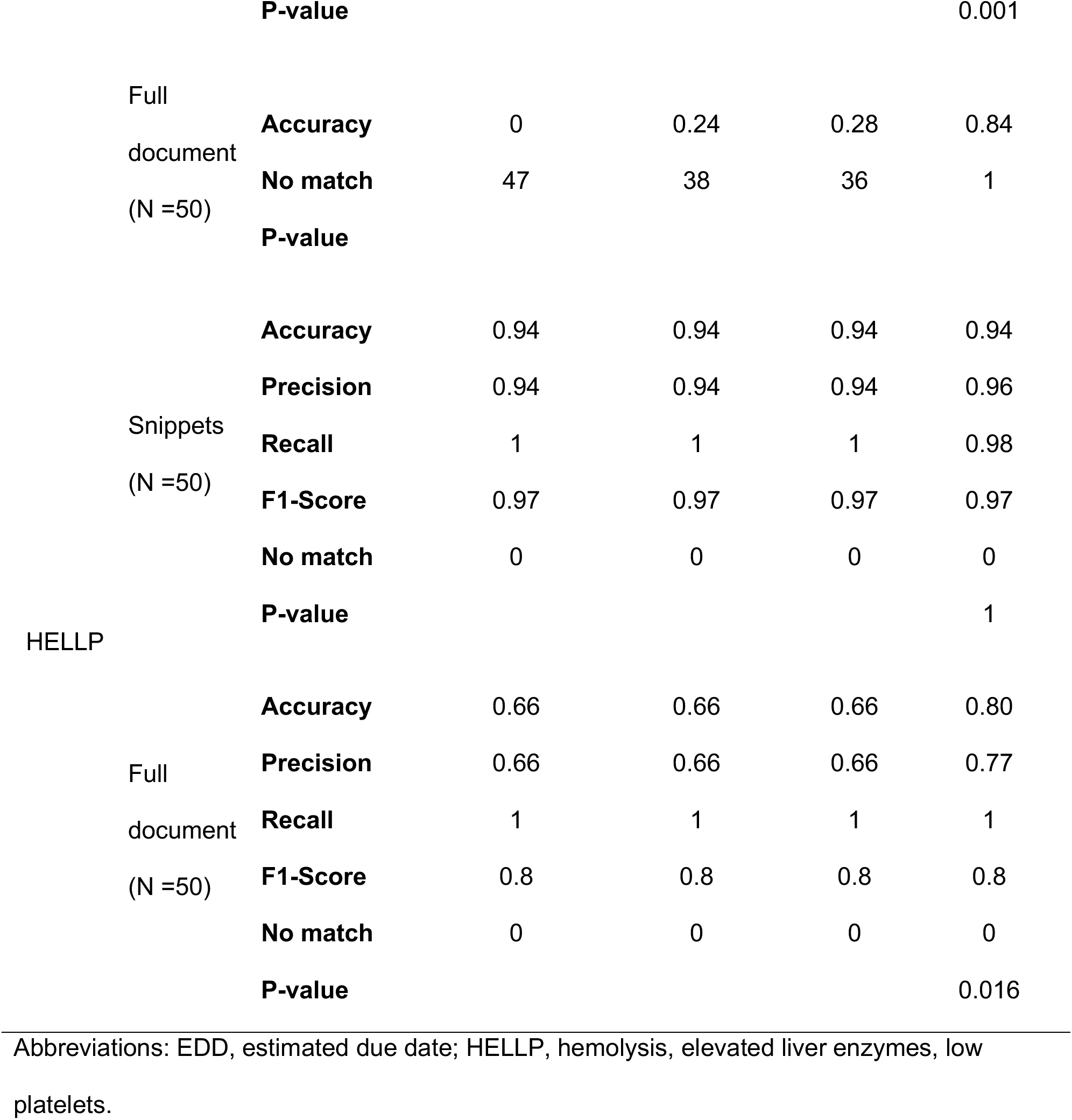
Performance metrics comparing regex-only baseline methods (Regex@N) versus LLM extraction. For each task and document subset (snippets vs. full document), P-values are from McNemar’s exact test on paired document-level correctness, comparing theregex-only baseline with a 5-word window to the LLM extraction. “No match” indicates that a method did not produce a candidate output for that document; all ‘No match’ cases were counted as incorrect when computing performance metrics.

The accuracy of the LLM-assisted extraction was rigorously evaluated against manual annotations (Table 3). For BL quantification, the LLM achieved 98% accuracy, outperforming retrieval-based baselines (Regex@1 = 22%), with no unmatched cases. For EDD extraction, the LLM reached 94% accuracy, exceeding all retrieval-based methods (Regex@1 = 20%). In the HELLP syndrome detection task, the LLM achieved 94% accuracy, 0.96 precision, 0.98 recall, and an F1-score of 0.97, with no unmatched cases.

When compared against annotations derived from complete-note datasets (i.e., cases where annotators labeled full documents rather than curated snippets), the LLM maintained strong performance: 94% accuracy for BL quantification, 84% for EDD, and 80% for HELLP, demonstrating robustness to annotation granularity. Across tasks, residual errors primarily stemmed from contextual ambiguities such as distinguishing current from prior pregnancies and overlapping temporal references within the clinical text.

By comparison, regex-only approaches demonstrated significantly lower reliability, underscoring the importance of the contextual interpretation capabilities of the LLM.

### 4.5 Computational Performance

Inference time scaled linearly with input length, demonstrating efficient computational resource use. GPU monitoring indicated stable, sustainable, and scalable hardware performance, making it suitable for institutional implementation (detailed GPU metrics are provided in Appendix S1). Performance greatly benefited from our snippet-based approach, allowing us to obtain a 5.57 ± 1.73 speedup on average across tasks.

### 4.6 Practical Workflow Example (EDD Extraction)

Figure 2 demonstrates a practical, step-by-step workflow example: regex-based note selection, snippet extraction, and structured data extraction via LLM inference. This illustrates the pipeline’s potential for intuitive integration into routine clinical informatics workflows (detailed description in Appendices S7–S9).

### 4.7 Generalizability Evaluation Across Clinical Contexts

To evaluate SPELL’s generalizability beyond obstetrics, we tested its performance in identifying ventricular tachycardia diagnoses in the MT Samples dataset. SPELL achieved high extraction accuracy (84%) and an F1-score of 67%, demonstrating robust performance despite substantial differences from obstetric documentation. The snippet-based inference strategy significantly enhanced efficiency, completing the task in 2.74 seconds compared to 15 minutes for full-document analysis, and yielded approximately 3.5-fold fewer false positives. These findings underscore SPELL’s adaptability and potential for broad clinical informatics applications.

## 5. DISCUSSION

In this study, we developed and validated SPELL, a scalable and modular NLP pipeline designed to automatically extract structured clinical insights from EHR narratives. The hybrid framework combined precise regex-based snippet extraction with locally hosted LLM inference, demonstrating substantial computational efficiency gains and high accuracy across three representative obstetric tasks: quantifying BL, retrieving EDD, and detecting HELLP syndrome. These results were further validated using a publicly available dataset.

### 5.1. Key findings and implications

Our results demonstrated substantial advantages of SPELL over both manual physician annotation and purely regex-based methods. Specifically, SPELL significantly reduced manual review effort, achieving 97% time savings compared to manual annotations. Furthermore, the platform consistently matched expert-level accuracy across diverse clinical extraction tasks, including validation with the MT Samples dataset, highlighting its potential to transform the feasibility of large-scale, retrospective clinical studies and routine clinical analytics [1,3,15].

A critical contribution of our approach was its structured workflow, modular architecture, and inherent flexibility to integrate emerging generative AI technologies seamlessly [13,15]. Unlike specialized NLP pipelines that typically require intensive manual annotation, feature engineering, and frequent model retraining [8,9,12], our platform was explicitly designed for rapid adaptability and ease of use. The modular strategy specifically anticipated ongoing advances in generative AI, enabling clinical informatics teams to harness clinical narrative data effectively without extensive technical expertise or investment in specialized infrastructure.

### 5.2. Computational efficiency and resource optimization

We implemented a dynamic, single-sample allocation strategy that addressed inefficiencies associated with traditional batching methods typically recommended for GPU optimization. In classical machine learning pipelines, batching inputs into fixed-length tensors is considered best practice for computational efficiency [33]. However, in clinical NLP, considerable variability in note lengths causes excessive padding and memory overhead, thus degrading efficiency [34]. By dynamically loading each snippet individually, our pipeline eliminated padding-induced computational bottlenecks, achieving higher processing speeds and more predictable linear scaling of inference time relative to input length. Our approach resulted in significant time savings when scaled to millions of clinical documents, reducing computational costs and substantially lowering GPU energy consumption (approximately a 9.2-fold reduction in cumulative energy usage for the same set of notes) and operating temperatures (by an average of 5.4 °C per GPU; Appendix S1), thus promoting hardware sustainability and operational longevity.

### 5.3. Novelty and differentiation from existing approaches

SPELL significantly differentiated itself from traditional NLP methodologies, including ontology-driven systems such as cTAKES and MetaMap [29,30] and transformer-based clinical models such as ClinicalBERT or BioBERT [23,24]. While ontology-based systems rely heavily on standardized vocabularies (e.g., UMLS, SNOMED CT) and manual maintenance [29,30], they may encounter performance limitations when extracting nuanced, rare, or context-dependent clinical concepts [4]. Similarly, transformer-based NER pipelines require substantial domain-specific fine-tuning and annotated datasets, which can increase operational complexity and limit rapid adaptability [22–24].

A recent study developed a similar regex-LLM framework tailored specifically for spinal surgery data extraction; however, it relied on cloud-based GPT-4 processing and performed inference at the full-document level, resulting in increased computational demands [18]. Another related approach followed a comparable annotation protocol but focused on a single type of clinical report and processed each line independently rather than leveraging regex-identified snippets, which led to notably smaller gains in annotation speed relative to ours [35]. Another recent method introduced clinical entity-augmented retrieval to enhance extraction efficiency and accuracy in clinical notes, but, while it reported inference-time and token-usage metrics, it did not quantify manual annotation burden or provide explicit evaluation across a broad range of clinical specialties [31]. Additionally, an evaluation of open-source LLMs demonstrated improved accuracy for extracting social determinants of health compared to traditional regex methods, though without prioritizing local hosting, explicit scalability testing, or comprehensive computational metrics [14].

In contrast, SPELL uniquely combines systematic regex-based snippet retrieval with locally hosted generative LLM inference, enabling high computational efficiency, accuracy, and zero-shot adaptability. Moreover, our pipeline explicitly restricts inference to targeted textual snippets, substantially reducing computational load, and uniquely includes detailed computational metrics such as GPU throughput, energy consumption, and optimized scheduling, demonstrating institutional-scale application across millions of clinical documents.

### 5.4. Potential clinical and operational impact

The substantial, 97%, reduction in manual annotation workload achieved by SPELL translated into significant operational improvements, potentially streamlining workflows in clinical research, population health management, and clinical quality assurance. Automating data extraction from clinical narratives not only freed clinicians from resource-intensive annotation tasks but also allowed greater focus on clinical interpretation, intervention planning, and patient care quality improvement initiatives. Furthermore, SPELL demonstrated flexibility and modularity, positioning it uniquely for supporting clinical decision-making through real-time or near-real-time extraction of clinical insights from narrative documentation, thereby potentially enhancing patient safety, outcome monitoring, and predictive analytics capabilities [1,3,5].

### 5.5. Strengths of the proposed approach

A key strength of our approach is the clear separation between regex-based snippet retrieval and LLM-based contextual interpretation. This design allowed domain experts to rapidly refine queries and prompts tailored to new clinical scenarios, significantly enhancing usability and reducing operational barriers. The local hosting and inference approach addressed some institutional data governance concerns, facilitating deployment in healthcare organizations [15,16]. Additionally, our explicit demonstration of generalizability using the MT Samples dataset provided clear evidence of SPELL’s adaptability beyond the original obstetric scenarios, highlighting the pipeline’s potential for immediate adoption across diverse clinical domains. Furthermore, SPELL has been optimized to efficiently process millions of notes, enabling practical deployment at scale in real-world healthcare settings.

### 5.6. Limitations and future directions

Despite clear feasibility and strong performance, several limitations and opportunities for future improvement remain. Our primary evaluations focused on obstetric scenarios, chosen intentionally for their standardized terminologies and relatively explicit documentation. Although initial validation for ventricular tachycardia demonstrates promising generalizability, broader assessment across additional medical specialties and varied documentation practices is essential.

Our current model choice (Llama 3.1-8B-Instruct) prioritized practical scalability and efficiency. Although this model was not specifically fine-tuned for clinical text, recent literature suggests that instruction-tuned general-purpose LLMs with appropriate prompting and moderate fine-tuning can match or exceed the performance of some domain-specific models, such as BERT-based architectures, on certain clinical extraction tasks, while maintaining practical inference efficiency [17]. Future studies might evaluate hybrid approaches that integrate domain-specialized models or representations (e.g., ClinicalBERT, BioBERT) with snippet-based LLM extraction, potentially further enhancing accuracy [23,24].

Our current framework does not explicitly map extracted concepts to standardized terminologies (e.g., ICD-10, SNOMED CT), potentially limiting immediate interoperability with clinical systems. Future iterations should integrate such mapping alongside dedicated handling of linguistic modifiers (negation, uncertainty) [36,37] to improve extraction reliability.

While our regex recall audits confirmed high retrieval coverage, systematic evaluation of regex robustness and comparison with alternative embedding-based or hybrid retrieval methods could further strengthen methodological generalizability. Additionally, our snippet-based inference approach, optimized for explicit clinical concepts, may face limitations with nuanced temporal reasoning or implicit documentation contexts. Exploring selective integration of long-context models to complement snippet-based extraction may offer valuable future directions.

Finally, external validation across diverse institutions, proactive monitoring of concept drift in evolving documentation practices, and demographic fairness analyses are critical next steps toward robust and equitable clinical NLP deployment.

### 5.7. Comparison with existing methodologies

While structured clinical data warehouses and structured query systems (e.g., i2b2) offer systematic extraction alternatives[32,38], these solutions require significant initial investment and infrastructure adjustments. In contrast, SPELL leveraged existing institutional EHR infrastructures and standard NLP libraries, providing immediate practical utility, significantly lower setup costs, and rapid deployment capabilities.

### 5.8. Practical considerations for clinical deployment

Clinician involvement is critical for defining extraction criteria, guiding prompt design, and interpreting ambiguous cases. Simplified guidance or automated parameter tuning (e.g., context window sizing, prompt refinement) could further enhance usability for clinical informatics teams without specialized NLP expertise. SPELL’s modular Python architecture, structured outputs, and locally hosted design facilitate future interoperability with EHR systems, supporting potential real-time clinical decision-making and improved patient-centered outcomes.

## 6. CONCLUSION

SPELL demonstrated strong potential as a robust and efficient NLP pipeline for automating information extraction from EHR narratives. Its modular, scalable design effectively addressed critical informatics challenges, significantly reducing annotation workloads, improving operational efficiency, and enhancing clinical research capabilities. Successful validation within obstetric scenarios demonstrated feasibility, establishing a clear foundation for extending this approach broadly across diverse clinical settings, thus supporting more effective clinical research and enhancing evidence-based decision-making in healthcare.

## Data Availability

In accordance with the Mass General Brigham Institutional Review Board requirements, the research data supporting this project may not be publicly shared. The MT Samples dataset is publicly available at www.mtsamples.com.

## Abbreviations

BL: blood loss
BWH: Brigham and Women’s Hospital
EDD: estimated due date
EHR: Electronic health records
GPU: graphics processing unit
HELLP: hemolysis, elevated liver enzymes, and low platelets
ICD: International Classification of Diseases
MGB: Mass General Brigham
MGH: Massachusetts General Hospital
NWH: Newton-Wellesley Hospital
WDH: Wentworth-Douglass Hospital
SRH: Spaulding Rehabilitation Hospital
FH: Brigham and Women’s Faulkner Hospital
NSM: North Shore Medical Center
NLP: Natural Language Processing
SPELL: Snippet-Primed rEgex LLM Pipeline
MEE: Mass Eye and Ear
LLM: Large Language Model
UMLS: Unified Medical Language System

## Declaration of Generative AI and AI-assisted Technologies in the Writing Process

During the preparation of this work, the authors used OpenAI ChatGPT (accessed September 2025) in order to assist with language polishing and consistency checks. After using this tool/service, the authors reviewed and edited the content as needed and take full responsibility for the content of the published article.

