## Supplemental File for "SPELL: A Scalable NLP Method Using Regular Expressions and Large Language Models for Clinical Information Extraction"

**ARTICLE TITLE**:

**SUPPLEMENTARY MATERIAL**

**Appendix S1: Detailed computational evaluation and accuracy metrics**

To rigorously assess performance, we measured information-extraction accuracy, and computational scalability across pipeline stages. Memory consumption was tracked for raw clinical notes, intermediate regex outputs, and LLM outputs. We also recorded elapsed time for dataset creation, regex-based retrieval, and LLM inference to characterize computational cost.

**Accuracy and statistical testing.** For all three tasks, BL, EDD, and HELLP, the primary endpoint was exact-match accuracy (correct vs. incorrect relative to adjudicated labels). Paired method comparisons between the LLM and the regex baseline (Regex@N) used McNemar’s exact test on paired accuracy outcomes. No-match outcomes (i.e., no candidate found) for the regex baseline are counted as incorrect and are also reported separately as ‘No match’.

**Efficiency and telemetry**. To contextualize efficiency, we compared elapsed time for automated methods with a trained physician’s manual chart review of the same snippets for a subset of notes. We computed energy efficiency as the total power (W) required in a process from power logs as defined in Methods 3.5, and GPU temperature (°C), during inference. GPU metrics indicated consistent power draw and temperatures within optimal operating ranges (Fig. S1), suggesting efficient hardware utilization, reduced thermal stress, increasing operative stability, and longer hardware lifespan compared to approaches that process entire clinical reports.

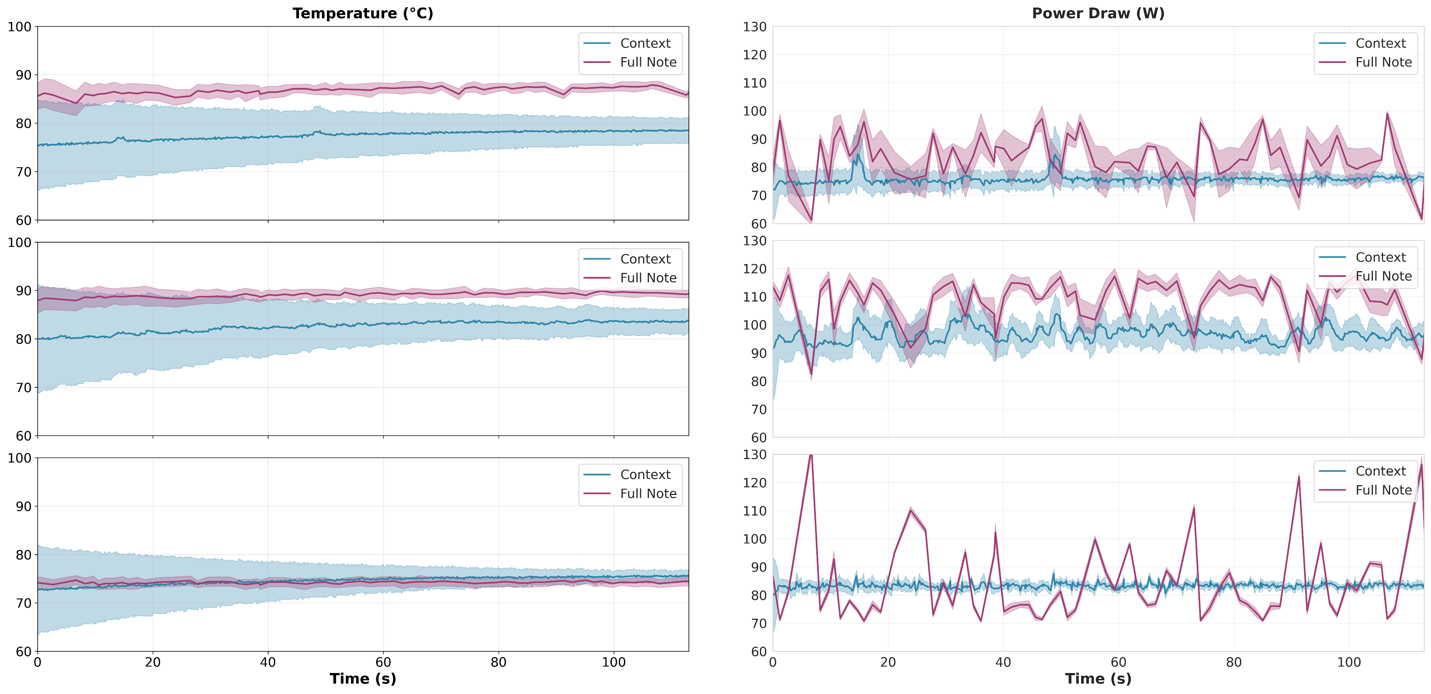

**Fig. S1.** GPU telemetry during inference, averaged across 10 repetitions, standard deviation signaling differences between them. Temperature and power, per GPU (one per row). The x-axis has been limited to match the shortest processing (snippet-based) to improve visualization.

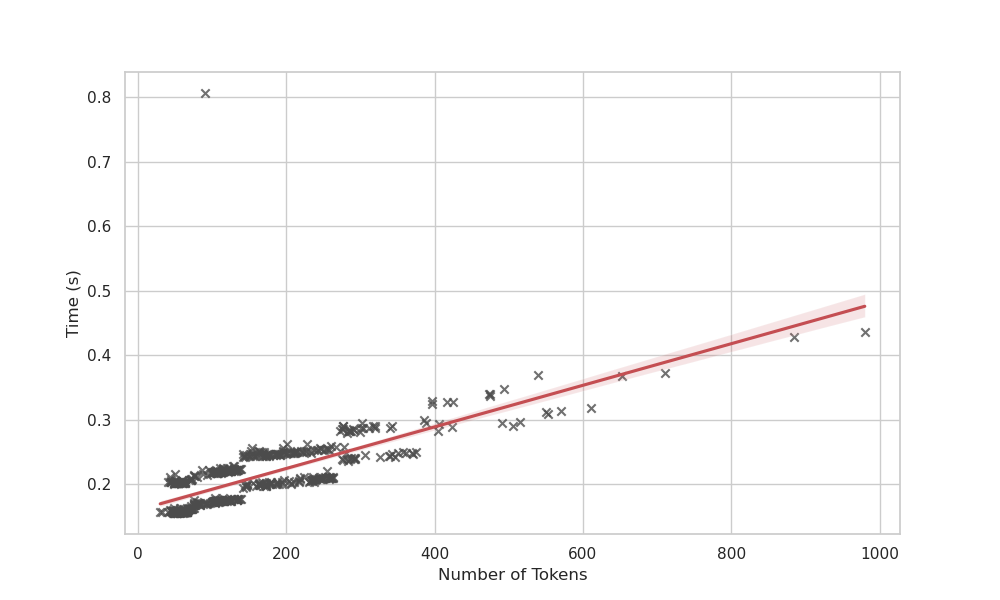

**Figure S2**. Elapsed time vs. input length. Elapsed time (seconds) as a function of tokens per sample. The first sample appears as an outlier (warm-up); thereafter, the relationship is approximately linear.

**Appendix S2: Number of patients per hospital for which data were available.**
Abbreviations: WDH, Wentworth-Douglass Hospital; SRH, Spaulding Rehabilitation Hospital; MEE, Mass Eye and Ear; FH, Brigham and Women’s Faulkner Hospital; NSM, North Shore Medical Center; NWH, Newton-Wellesley Hospital; MGH, Massachusetts General Hospital; BWH, Brigham and Women’s Hospital.

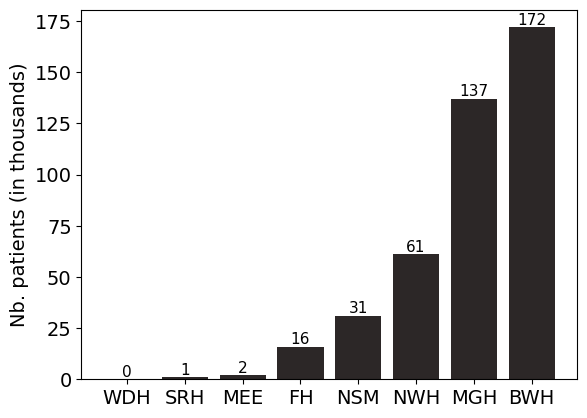

**Appendix S3:** Number of clinical reports available per year in the files downloaded from the data warehouse service as of the download date. Dates span March 1, 1976, to July 15, 2024.
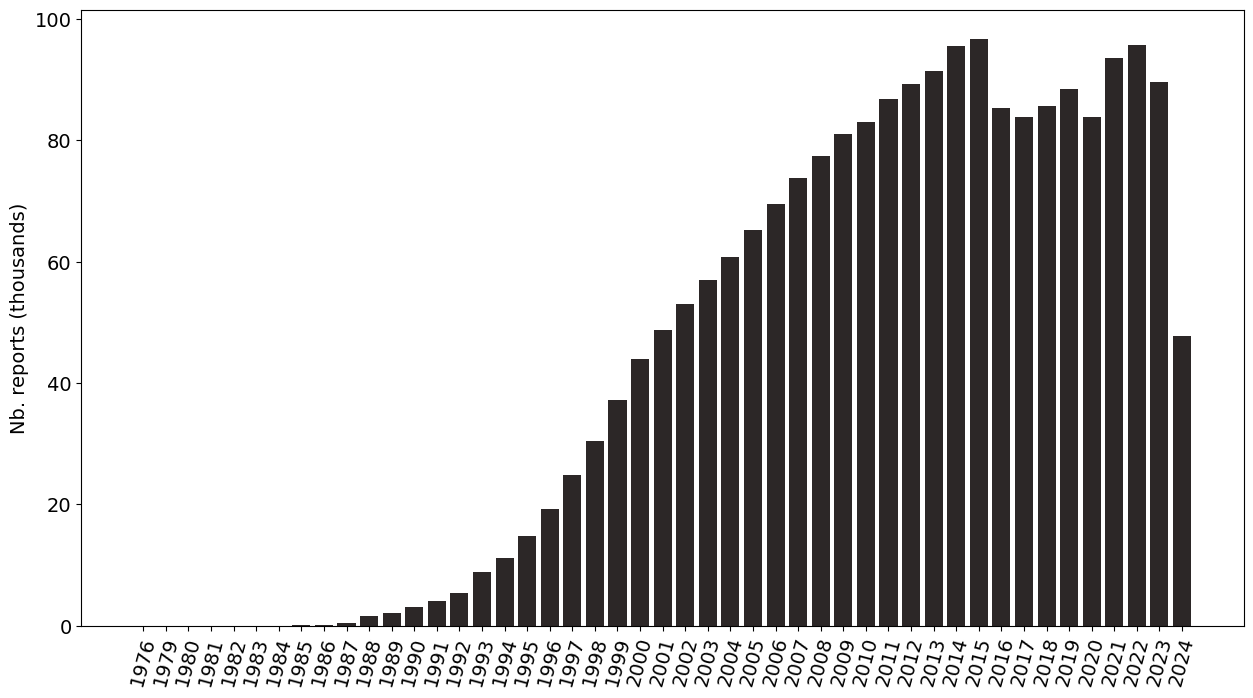

**Appendix S4:** Number of reports per type of note.

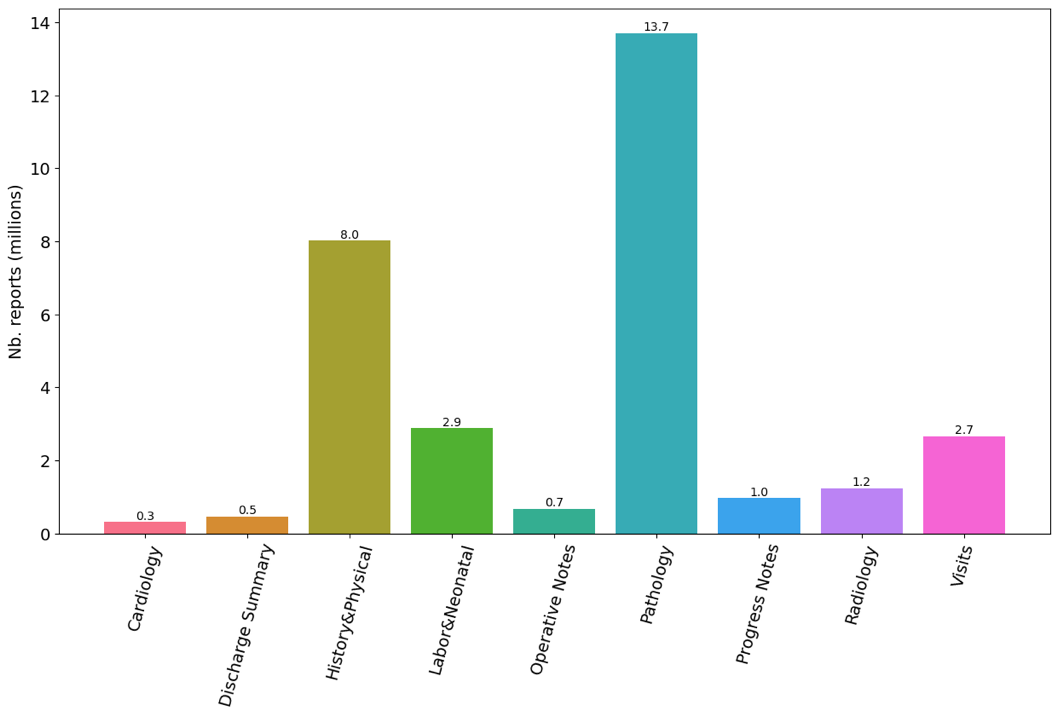

**Appendix S5:** Average number of words (mean ± SD) in the clinical reports contained in the files downloaded from the data warehouse, per report class.

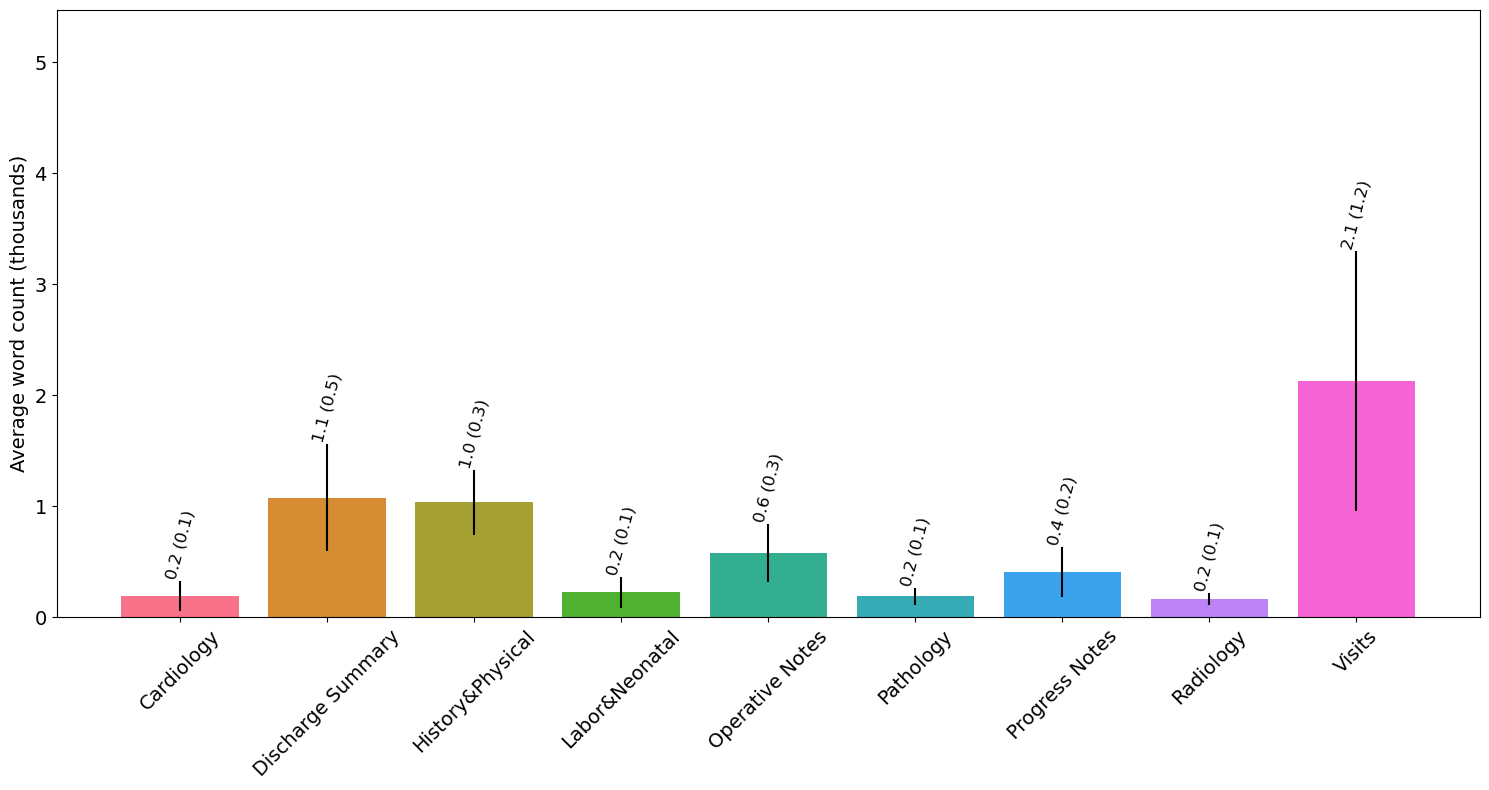

**Appendix S6:** Regular expressions considered for snippet extraction

| **Task** | **Use case** | **Regular expression** |
| --- | --- | --- |
| Boolean detection | HELLP syndrome | (?i)\bH\.?E\.?L\.?L\.?P\.?\b |
| Datetime extraction | Estimated Due Date (EDD) | (?i)\b(due\s*date\|EDD\s*-?\s*expected\s*date\s*of\s*confinement\|EDD\s*-?\s*expected\s*date\s*of\s*delivery\|estimated\s*date\s*of\s*(birth\|confinement\|delivery)\|expected\s*date\s*of\s*confinement)\b |
| Numerical value retrieval | Blood loss in milliliters | (?i)\b(estimated\s*blood\s*loss\|quantitative\s*blood\s*loss\|blood\s*loss\|q\.?b\.?l\.?\|e\.?b\.?l\.?)\b |
| *Case-insensitive matching; extraction windows default to ±100 characters. Date normalization enforces YYYY-MM-DD; units normalized to mL (e.g., ‘1 L’→1000 mL; ‘cc’→mL).* | | |

**Appendix S7:** Data selection for obstetrics-related extraction tasks

To evaluate both extraction quality and computational efficiency, we applied additional constraints to limit the size of the datasets used in obstetrics-related extraction tasks. In all cases, only 147,496 pregnancies corresponding to 110,434 patients presenting from 2015 onward were considered, ensuring consistency in documentation practices.

For the Blood Loss (BL) extraction task, we included only obstetric (OB) discharge summary notes. Postpartum OB discharge summary notes were unavailable for 10,838 patients (9.81%), and these cases were excluded. Additionally, for another 6,824 patients we could not obtain a positive regex-based match for this task, so we excluded them from the study as well. For the remaining pregnancies, we restricted the dataset to notes written within the 48-hour period before delivery up to 60 days postpartum, resulting in a final dataset of 92,380 OB discharge summaries.

For the Estimated Due Date (EDD) extraction task, we considered the same initial cohort of patients but restricted the dataset to History & Physical (H&P) notes recorded between the estimated conception date and the date of delivery. 14,163 patients (12.82%) were excluded because we could not find any H&P notes. Remaining pregnancies with at least one note from which snippets could be retrieved amounted to a total final dataset size of 35,172 H&P notes.

For the HELLP Syndrome detection task, we focused on OB discharge summary notes from 1,776 pregnancies involving 1,701 unique patients with a positive HELLP Syndrome diagnosis based on registered ICD codes. OB discharge summaries were unavailable for 120 pregnancies (6.76%), and these cases were excluded. Applying the same selection criteria as in the BL task, we retained 540 OB discharge summaries written within the 48-hour period before delivery up to 60 days postpartum.

**Appendix S8:** Prompt templates

Every note, or list of contexts extracted for a given set of regex detections, is formatted according to the following template, which, for the case of Llama3.1-8B-Instruct, includes a set of system instructions that tailor the overall tone and constraints of the responses. The complete template used throughout our experiments is the following:

You are an expert physician. You are going to be presented with segments of clinical reports from pregnant patients separated by `|`, and your task is to retrieve the piece of information I will ask you about. Do not provide any explanation, simply return the requested information. Do not make up any information if you can't find it. The clinical report is:

<<snippets>>

[Question (task-dependent)]

Where the questions for the tasks considered are:

HELLP: Is there a confirmed diagnosis of HELLP syndrome, or a strong suspicion from the physician, in this report, for the current pregnancy? Answer yes or no.

EDD: "What is the estimated due date (EDD)? If mentioned in the note, provide the date in the format YYYY-MM-DD, else provide None."

Blood loss: What is the total blood lost in milliliters? Sometimes, we use abbreviations like quantitative blood loss (QBL) or estimated blood loss (EBL). If mentioned in the note, extract the value, if not, extract none. If there is more than one value, extract all.

**Appendix S9:** Tokenized snippet length (by task).

| **N Tokens** | **HELLP (N =540)** | **EDD (N = 35,172)** | **Blood loss (N = 92,380)** |
| --- | --- | --- | --- |
| Mean | 155.06 | 94.95 | 209.77 |
| SD | 118.46 | 66.58 | 73.65 |
| Min | 30 | 17 | 37 |
| Q1 | 66 | 60 | 181 |
| Median | 117 | 68 | 198 |
| Q3 | 206 | 114 | 258 |
| Max | 980 | 1,119 | 1,638 |
| *Tokenizer: Llama 3.1 byte-level Byte-Pair-Encoding tokenizer.* | | | |
